# EFFECT OF DIETARY CALCIUM ON INCIDENCE OF CALCIUM STONES AND URINARY PARAMETERS: A SYSTEMATIC REVIEW

**DOI:** 10.1101/2023.06.14.23291414

**Authors:** Dev Desai, Tanvi Sahni, Dev Umangbhai Andharia, Dwija Raval

## Abstract

**Introduction:** Renal stone is a fairly common presentation and the pain is unbearable. Some patients have tendency that stones are frequently formed and management of that s necessary as removal of stone is nt always that simple and can lead to many complications.

**Aims and Objectives:** To perform a systematic review and meta-analysis to investigate the effect of dietary calcium on the prevention of calcium stones.

**Methodology:** PubMed, google scholar and Cochrane Library databases were searched for studies. The search strategy and study selection process was conducted by following the PRISMA statement. 16 articles were identified after meeting the inclusion criteria.

**Results:** Our results showed that with high calcium advice, occurrence of stone formation decreases (RR=0.82, CI95=0.37-1.82, p=0.63). Comparing levels of Urinary caclium with advice of high calcium diet showed increased excretion (SMD=1.99), but decreased excretion with low calcium diet (SMD = -0.43) and Normal calcium diet (SMD = -1.35). An almost no difference can be seen in Urinary oxalate with high calcium (SMD = 0.00), low calcium (SMD = 0.02), Normal Calcium (SMD = -0.052). Results showed that Urinary Urate levels decreased with all types of diet mainly with High calcium diet (SMD = -0.33), then Low calcium diet (SMD = -0.28)and then normal calcium (SMD = -0.22). Urinary Volume was increased with all dietary advice, the most with low calcium diet(SMD = 216.37), then normal calcium diet (SMD = 206.81) and last high calcium diet (SMD = 20.00).

**Conclusion:** ur study shows high calcium diet decreases the occurrence of stones compared to normal calcium and low calcium diet.

## Introduction

Renal stone disease,also referred as Nephrolithiasis or Urolithiasis was first documented by the Egyptians.Even though the Management of kidney stones has evolved from open surgical lithotomy to minimally invasive endourological treatments over the decades, It still remains a disorder of the population are severely affected by renal dystrophy leading to loss of significant morbidity with high health care cost.

Prevalence of the Kidney stone disease shows variation by geography, mean age, dietary habits, fluid intakes etc. prevalence of the disease is 1 – 5% in Asia, 5 – 9% in Europe, 13% in North America, 4% in Argentina, 5.5% in Mexico and 20% in Saudi Arabia. [1] [2] [3] [4]. In the context of India,Kidney stone disease is prevalent, with an expectancy of 12%.Out of which, 50% of kidneys. [5] The annual cost of KSD in 1993 was $1.83 billion in the United States which increased to $2.07 billion in 2004 and is expected to reach $4.1 billion in 2030. [6]

The rising rate is attributable to changes in lifestyle and diet, resulting in increased obesity among men and women, a known risk factor for stone formation [7]. hypercalciuria is an important and common risk factor for the formation of stones, and uncontrolled hypercalciuria is a main cause of recurrences. [8]

Borghi et al.,RCT shows a positive impact of lower calcium diet in decreasing the stone occurnce rate where 23/60 from the control group got stone recurrence but 11/60 got recurrence from low calcium diet group [8] we wish to find a dietary solution,which can decrease the prevalence and battle the high cost of treatment and complications of KSD with the help of primary and secondary prevention.

## Methodology

### SEARCH STRATEGY

A search was done using the databases-PubMed,Google Scholar and cochrane library for all relevant literature. Full - Text Articles written only in English were considered.

The medical subject headings (MeSH) and keywords “Dietary advice”, “Calcium stone” and “Urinary parameters” were used. The last search was done on May 26th 2022. References, reviews and meta-analysis were scanned for additional articles.

### STUDY SELECTION

Titles and abstracts were screened, Duplicates and citations were removed. References of relevant papers were reviewed for possible additional papers.. Papers with detailed patient information and statically supported results were selected.

we searched for papers that compared diet advice with or without calcium, associated incidence of calcium stone and urinary parameters.

The inclusion criteria were as follows:; (1) studies that provided information about stone formation outcome; (2) studies published in English; (3) studies with statistical heterogenicity in terms of patient’s age and geography.

The exclusion criteria were as follows: (1) articles that were not full text ; (2) unpublished articles.

### DATA EXTRCTION

Each qualifying paper was independently evaluated by two reviewers. Each article was analysed for the number of patients, their age, dietary advice, incidence of calcium stone and urinary parameters like urinary calcium, urinary oxalate, urinary urate levels and total volumn of urine. Further discussion or consultation with the author and a third party was used to resolve conflicts. The study’s quality was assessed using the modified Jadad score.

### STATISTICAL ANALYSIS

All of the data was obtained and entered into analytic software. Fixed-or random-effects models were used to estimate mean difference, standardised mean difference (SMD), odds ratios, and relative risk (RR) with 95 percent confidence intervals to examine critical clinical outcomes (CIs). Statistical heterogeneity was measured with the χ^2^; P < 0.100 was considered as a representation of significant difference. I^2^ greater than or equal to 50% indicated the presence of heterogeneity. Funnel plots were used to assess potential publication bias based on the prevalence of wound infection after surgery. A statistically significant difference was defined as P<0.05.

## Results

### STUDY SELECTION

A total of 54,848 papers were generated from pubmed, google scholar and cochrane library. 32,280 papers were eliminated after not meeting the initial search criteria by title, 22,568 abstracts were selected for screening. Out of the 329, 313 were removed based on exclusion criteria; 13 were not available in English and the other 300 were not full text. The remaining 16 eligible papers were selected. 1 additional paper was retrieved from references. A total of 12 papers were selected for analysis. Flow chart 1 summaries the selection process.

### STUDY CHARACTERISTICS

The study characteristics of the studies are given in table 1. These studies investigated a total of 5700 patients ; who followed a high calcium diet, normal calcium diet and low calcium diet. Chart 2 depicts the effect of high calcium diet on incidence of kidney stones.

**Chart 2:**
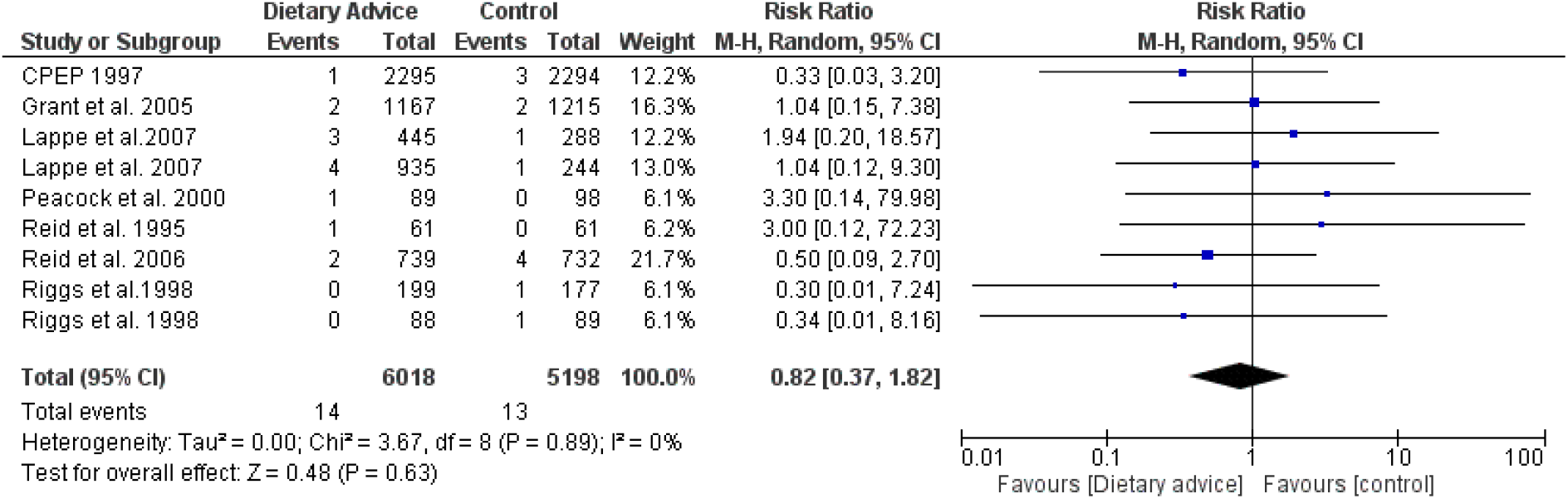
Effect of high calcium diet on incidence of kidney stones.

A random-effects model was used for analyzing the primary outcomes of kidney stone incidence. Our result showed that dietary intervention does decrease the occurrence of stone upon comparing with control groups (RR = 0.82, 95% CI = 0.37–1.82; P = 0.63). Borghi et al shows that a diet with a normal amount of calcium but with reduced amounts of animal protein and salt is more effective than the traditional low-calcium diet in reducing the risk of recurrent stones in men with idiopathic hypercalciuria

Urinary calcium excretion was taken as parameter in all the research papers and Dietary advice (DA) was given as a diet containing high calcium, low calcium,normal calcium. As shown in figure 1.1 High calcium diet is not advisable (SMD=1.99, C95=0.57-3.41, p=0.006). Figure 1.2 shows DA low calcium diet and shows heterogeneity (SMD=-0.43, C95=(−6.21)-5.34, p=0.88). Figure 1.3 shows DA as normal calcium (SMD=-1.35, C95=(−4.09)-1.38, p=0.33).

**Figure 1:**
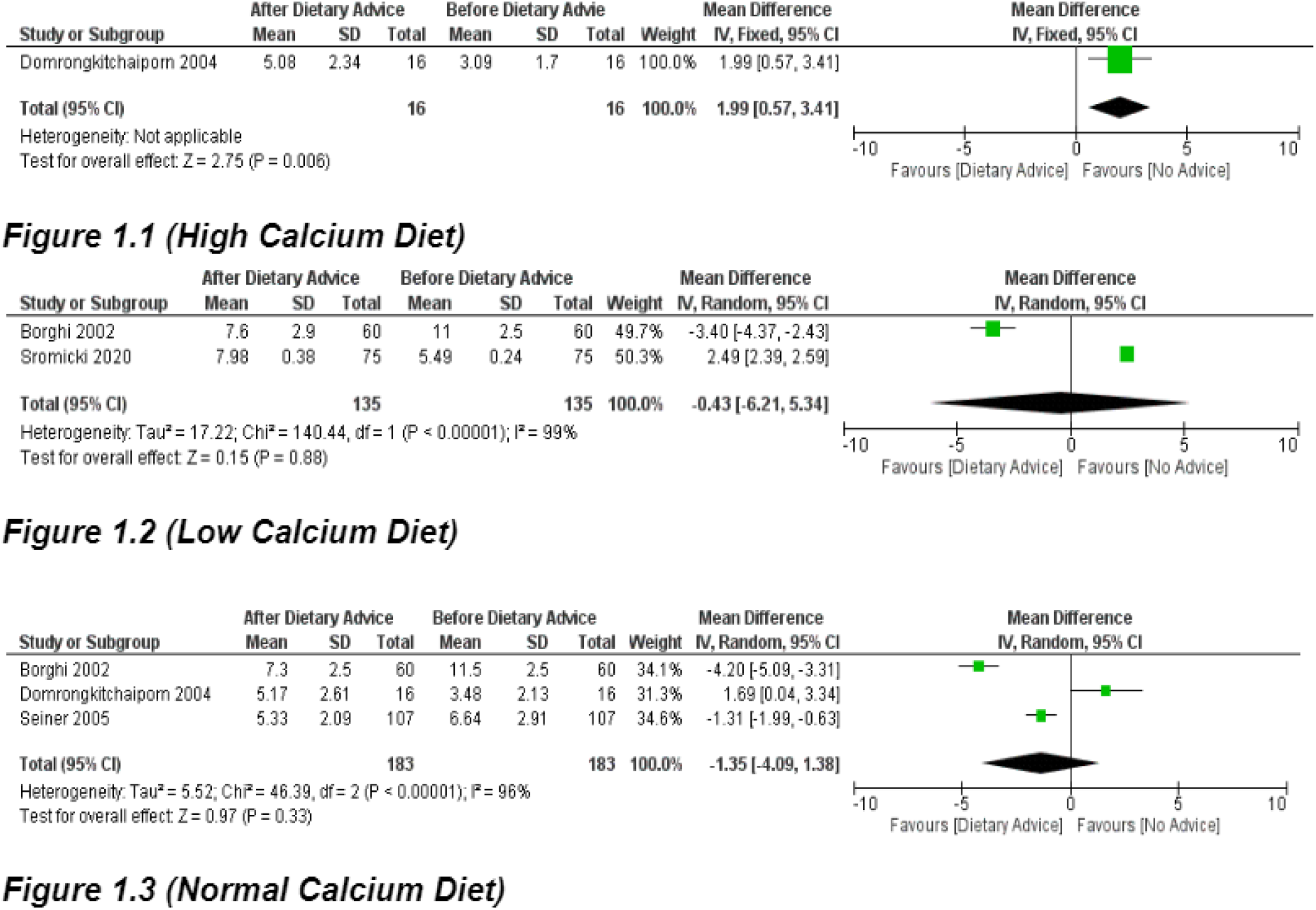
Urinary calcium excretion for high calcium, low calcium and normal calcium diet

**Figure 2:**
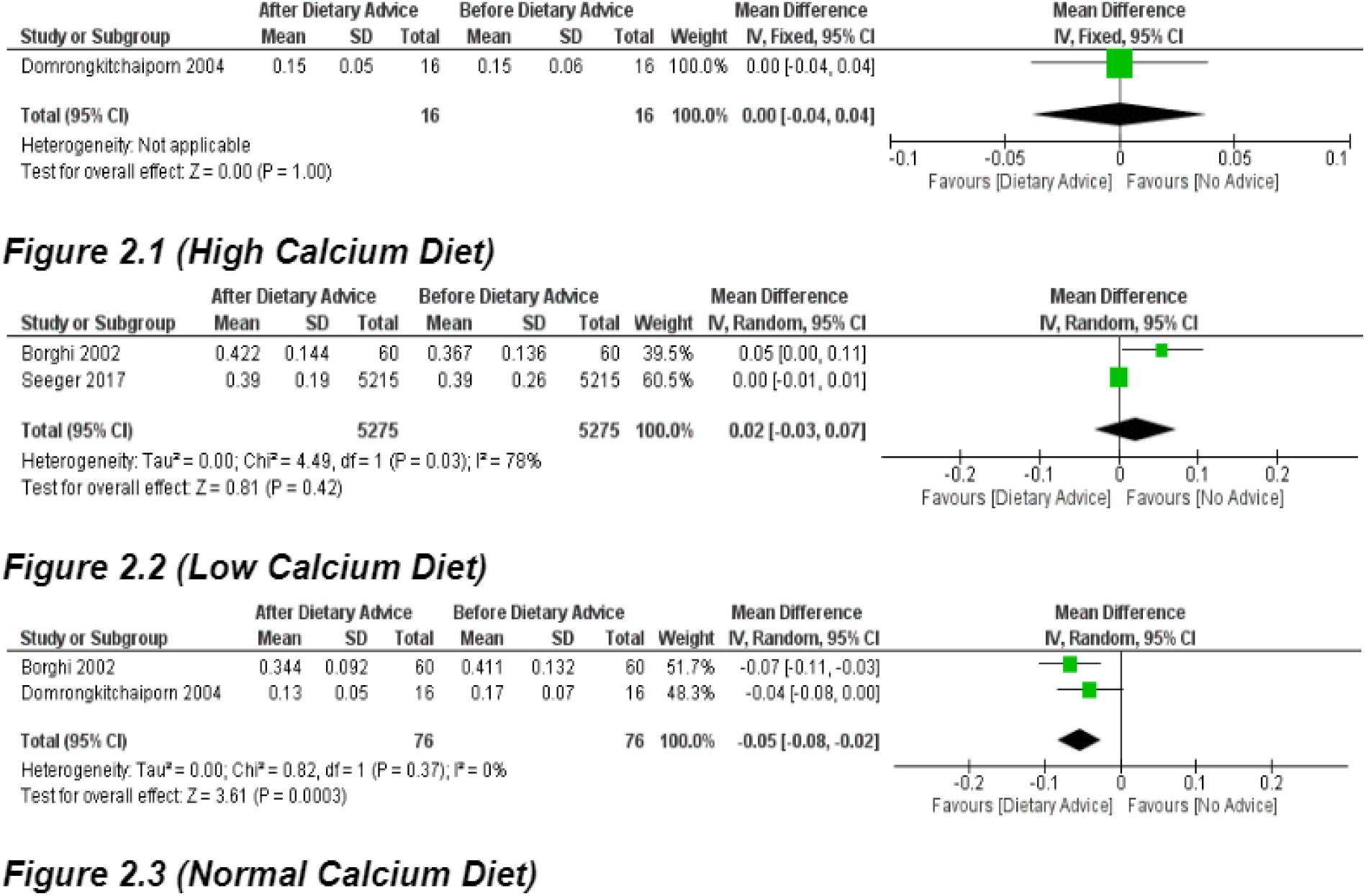
Urinary oxalate excretion for high calcium, low calcium and normal calcium diet

**Figure 3:**
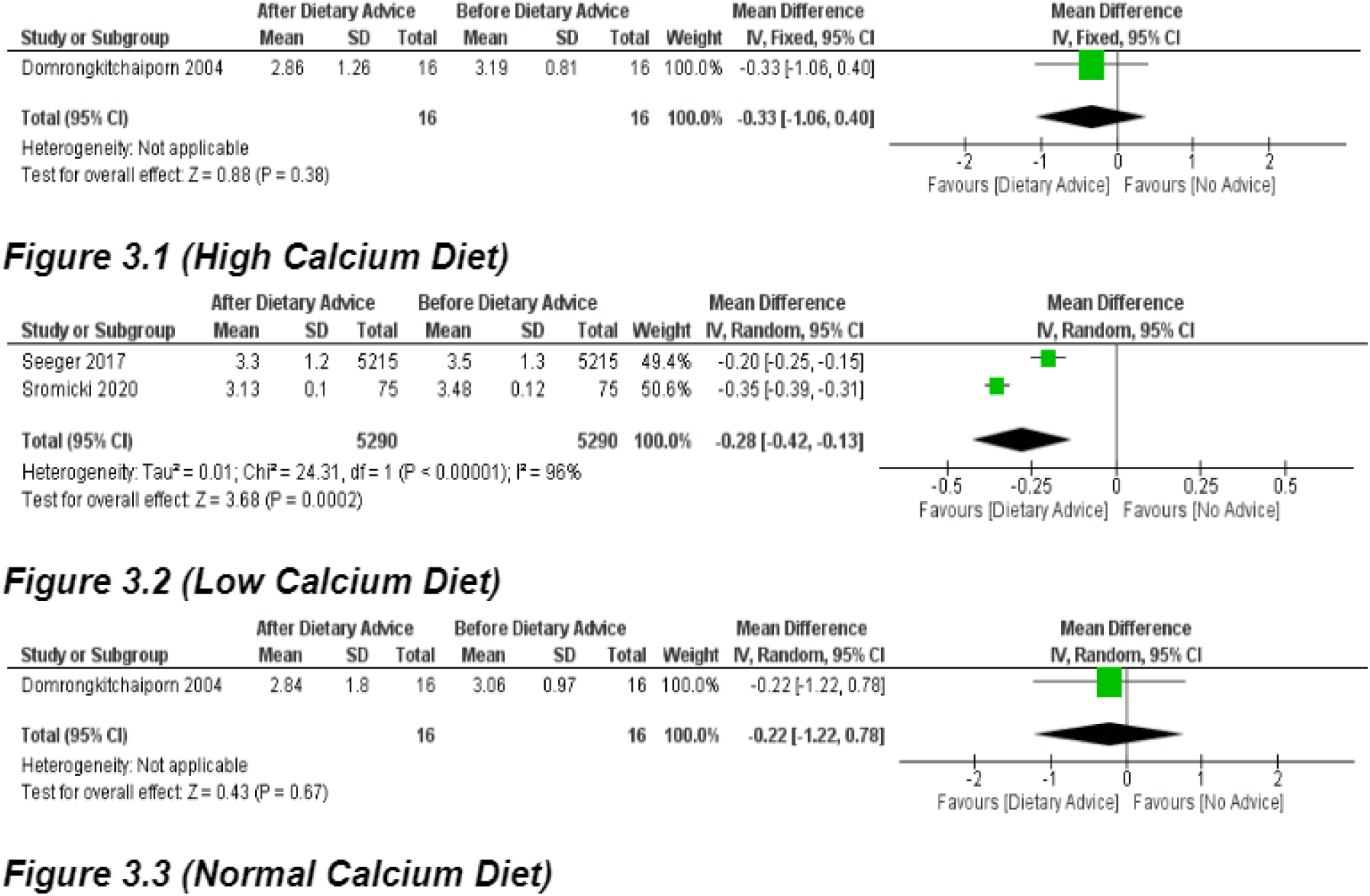
Urinary urate excretion for high calcium, low calcium and normal calcium diet

The urinary parameter is oxalate in Figure 2 with DA containing high calcium, low calcium, normal calcium. Figure 2.1 shows a high calcium diet (SMD=0.00, C95=(−0.04)-0.04, p=1.00). Figure 2.2 shows that low calcium diet (SMD=0.02, C95=(−0.03)-0.07, p=0.42).Figure 2.3 shows normal calcium diet (SMD=-0.05, C95=(−0.08)-(−0.02), p=0.0003).

The urinary parameter is Urinary urate in Figure 3 with DA containing high calcium, low calcium, normal calcium.Figure 3.1 is High calcium (SMD=-0.33, C95=(−1.06)-0.40, p=0.38). Figure 3.2 is low calcium diet (SMD=-0.28, C95=(−0.42)-(−0.13), p=0.0002).Figure 3.3 normal calcium is (SMD=-0.22, C95=(−1.22)-0.78, p=0.67).

The urinary parameter is Urinary volume in Figure 4 with DA containing high calcium, low calcium, normal calcium.Figure 4.1 shows high calcium diet (SMD=20.00, C95=(−289.84)-329.94, p=0.90). Figure 4.2 is a low calcium diet and (SMD=216.37, C95=(−223.21)-655.94, p=0.33). Figure 4.3 is a normal calcium diet (SMD=206.81, C95=(−7.06)-420.69, p=1.00).

**Figure 4:**
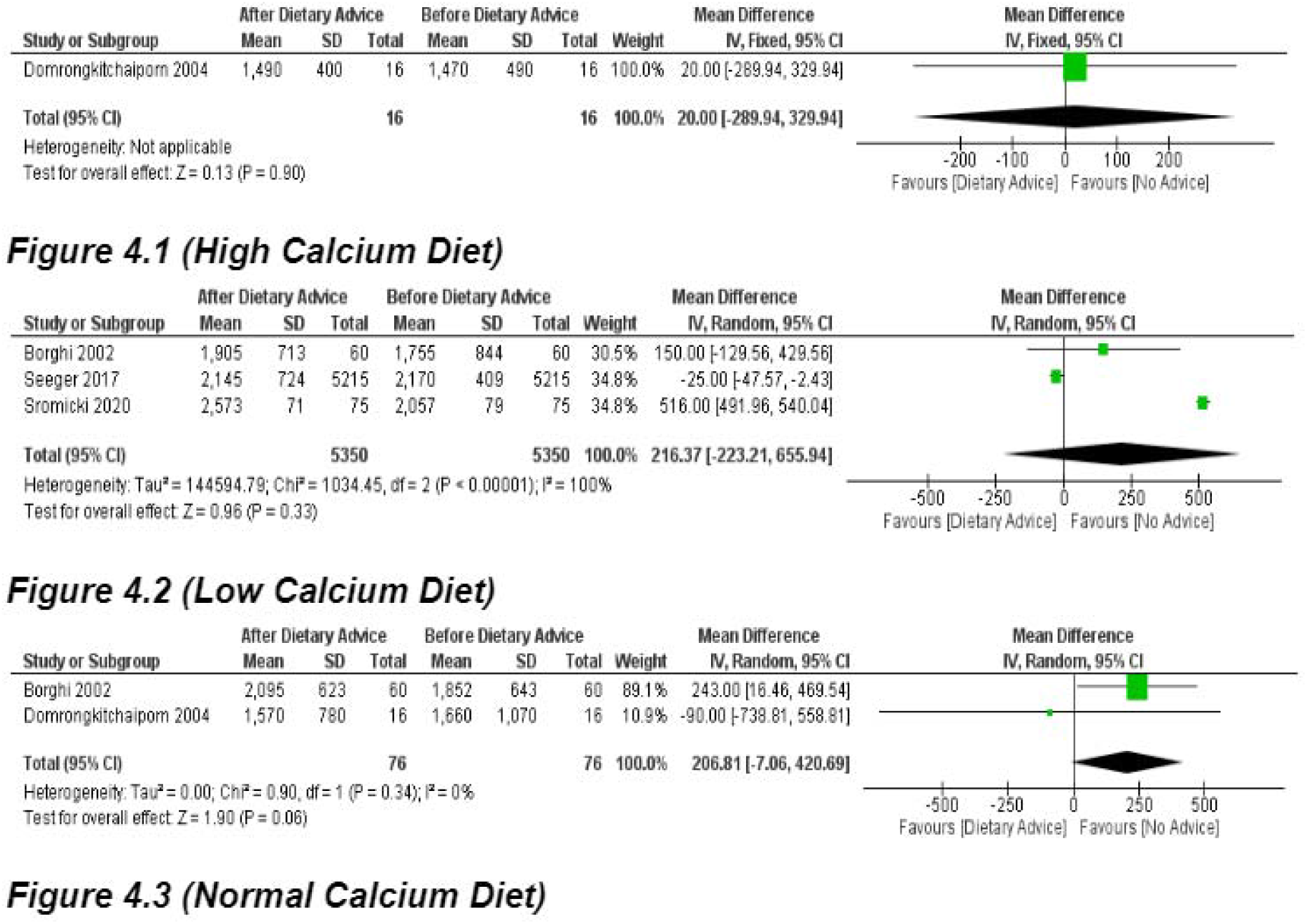
Urinary volume for high calcium, low calcium and normal calcium diet

## Discussion

Dietary calcium has been long associated with the of occurrence of renal stone and use of high dietary calcium diet has been identified as a protective factor in decreasing the incidence of renal stone formations. [9] [10] [11] it can be seen from these papers that high calcium diet advice should be given to individuals prone to developing renal stones, but it is also found by some papers that high calcium diet is actually a risk factor. [12] [13] [14] our results suggest that high calcium diet advice in individuals prone to developing calcium stone is actually protective and the incidence of developing renal stone decreases.

Total of 4 Urinary Parameters were considered which were Urinary Calcium, Urinary oxalate, Urinary Urate and total urine volume for each day. All these parameters were assessed with the different dietary changes and modifications to see changes in them and those papers were combined in this systematic review to find out the best dietary advice to reach target urinary parameters which leads to least calcium oxalate stone incidence.

The Urinary Calcium levels before and after dietary advice, as seen in figure 1 with a high calcium diet shows high urinary calcium excretion level which increases the risk of Calcium oxalate stone. [15] Although this is in line with common belief, Medical literature suggest that High Calcium diet advice should be given in recurrent stone formations as it decreases the level of urinary Calcium level. Only one paper where high calcium diet advice was given was found with sufficient data which can be the reason behind the result. In fact, what everyone believes is that a low calcium diet is a protective factor. [8] [16]

It is found to be in a lower efficacious protective role than Normal calcium diet. [8] [15] [17] The trend from low calcium diet and normal calcium diet suggest that high calcium diet should be a bigger protective factor than normal calcium diet advice.

In urinary oxalate levels before and after dietary advice as seen in figure 2 that high calcium diets do not have a role in urinary oxalate excretion. meanwhile it shows that dietary high calcium is neither positive nor negative factor for calcium oxalate stone formation. [15] As there is common belief that a low calcium diet could be protective in calcium oxalate stone formation we reviewed two papers suggesting that a low calcium diet is not a protective factor.

[18] [8] Hence it is not advisable to give a low calcium diet. However, the Normal calcium diet shows protective results against oxalate stone formation. [8] [15] Hence, Patients of Urinary oxalate stones are advised to take normal calcium in their diet rather than low or high calcium diet.

In Urinary urate levels before and after dietary advice are seen as in figure that high calcium diet is protective in urinary urate excretion. [15]. In a low calcium diet, 2 papers show that it is favourable to take low calcium in urate stones of the kidney. [18] [16]. Normal calcium is considered protective for urate stones. [15]. We observed that dietary calcium advice is minimal effective on urinary urate excretion.s.Hence we observe that the number of studies should be increased regarding calcium as dietary advice for urate stones.

One of the most effective methods for prevention of stone recurrence is increasing the urine output (>2 L/day)t.we measured urinary output (figure 4) before and after dietary advice. In a high calcium diet urinary output decreases but it is insignificant. [15]

In a low calcium diet, one paper with high sample size (approx 5000) shows that urinary output increases while other 2 papers with relatively low sample size show decreased urinary output. [8] [18] [16]. In a normal calcium diet urinary output is decreased and standard deviation is very high hence we can not conclude. [15]

The hypothesis behind decreased renal stone with high calcium diet could be that with high calcium intake, the body does not have to resorp the calcium and phosphate from the bones to maintain a steady blood calcium levels which means that excess calcium in the blood wont be excreted out through urine. And as high dietary calcium is being supplied, body will only absorb the amount of calcium it needs to not hamper the bone but also keep a steady blood leveles leading to less chances of stone formation in urinary tract.

After all this research we conclude that there is a lack of data and the number of studies are minimal, hence we should do more research regarding this subject to provide more proper knowledge of dietary advice in idiopathic recurrent stone formers.

## Conclusion

It can be understood here that dietary advice when given to consume high calcium diet, it decreases the occurrence of renal stone. The effect of different amount of calcium diet was found to be statistically insignificant on urinary parameters, although not many research papers are present to understand the effect of dietary advice on these urinary parameters properly. Hence, more prospective cohort studies should be carried out to find out the exact effect of the role of diet in preventing formation of stones.

## Data Availability

All data produced in the present work are contained in the manuscript

## Description of papers

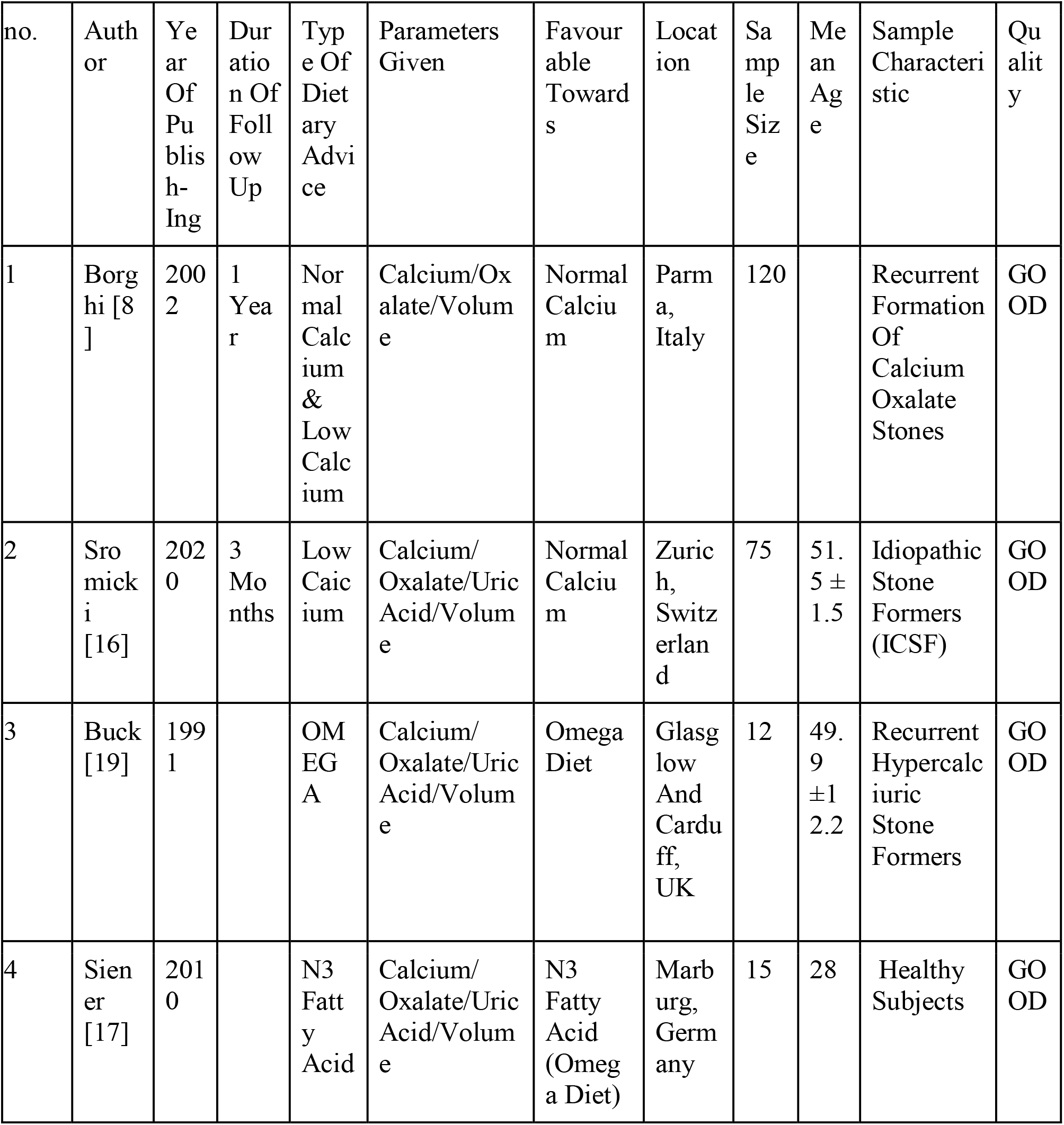
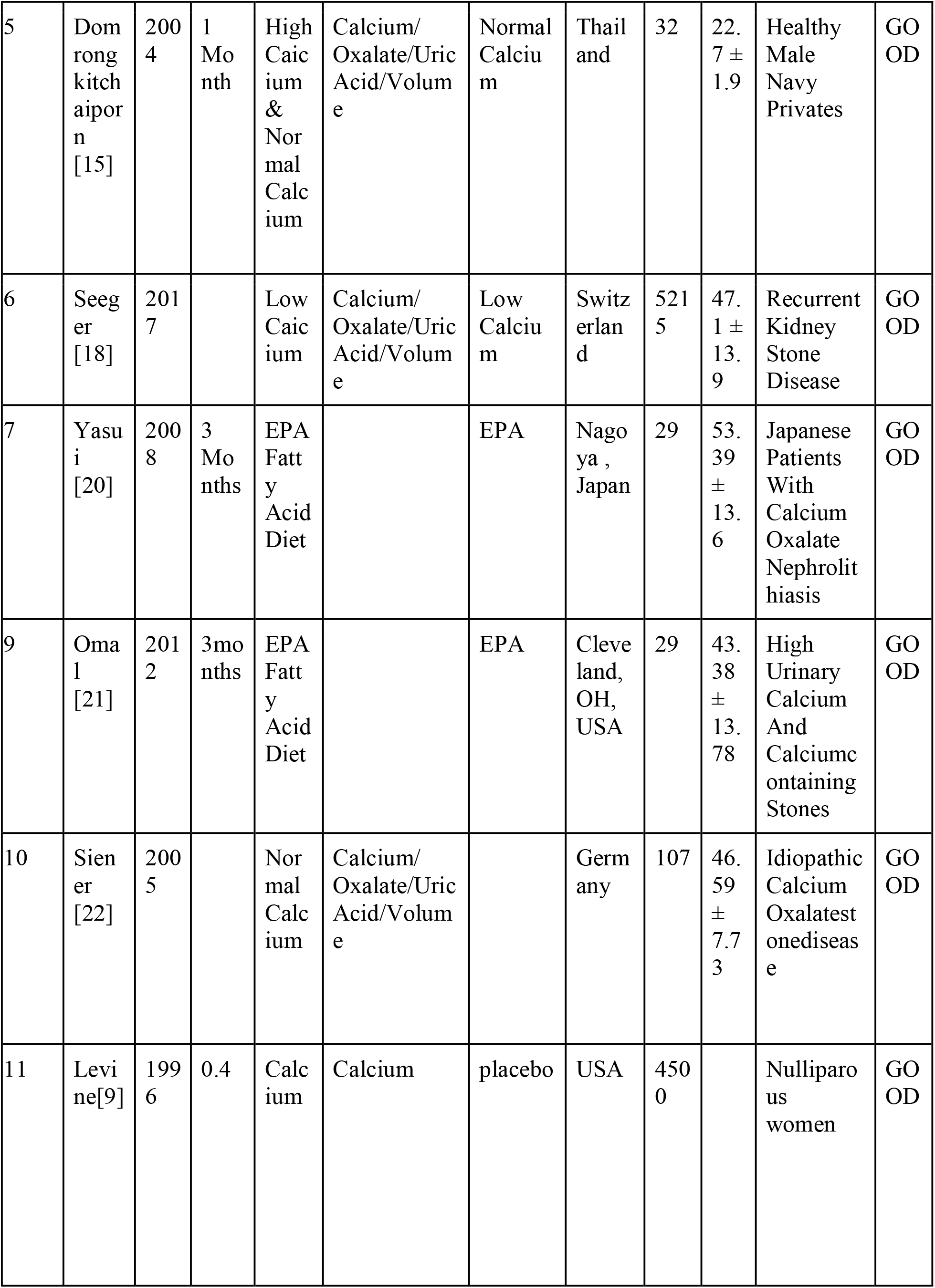
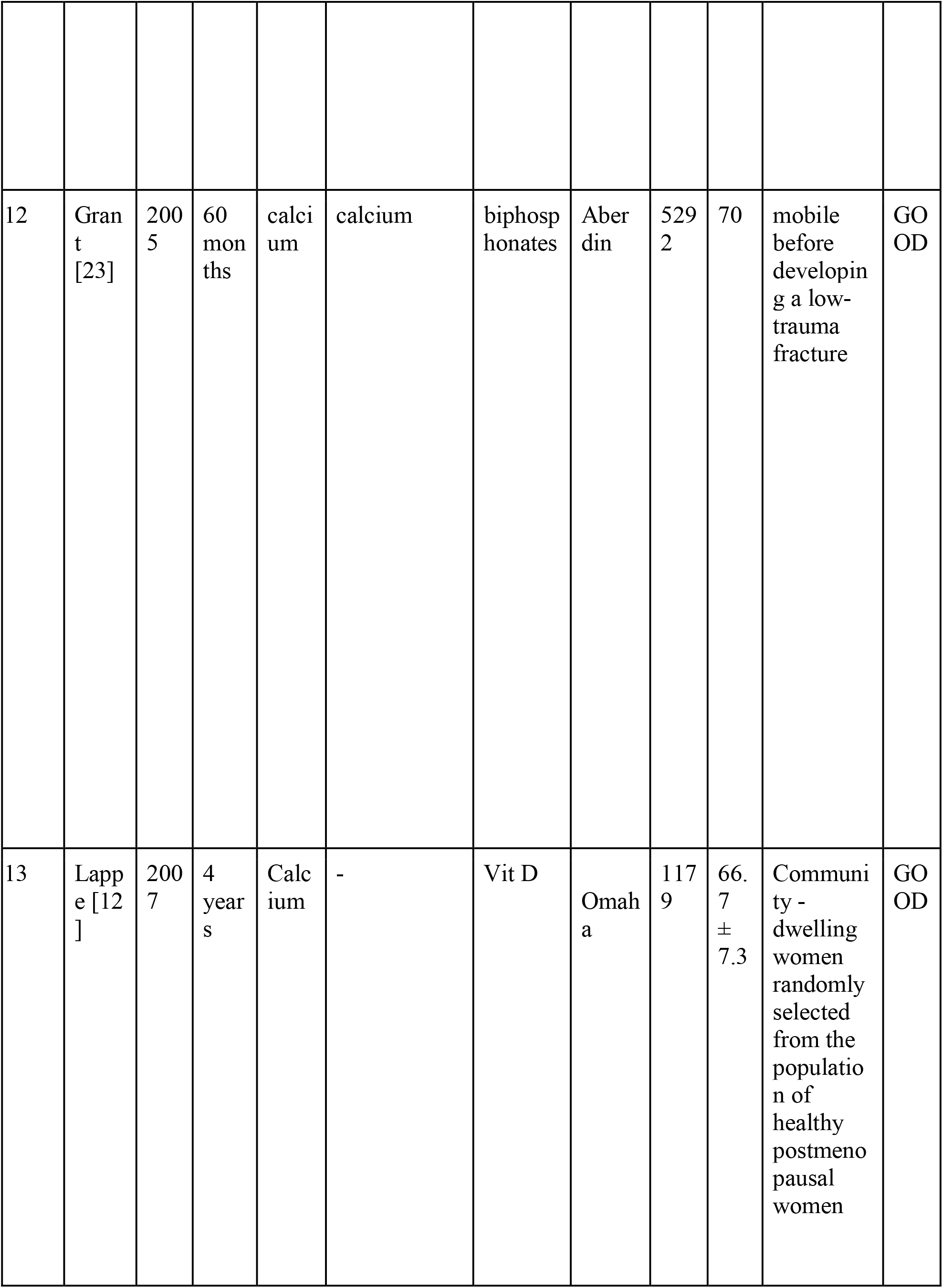
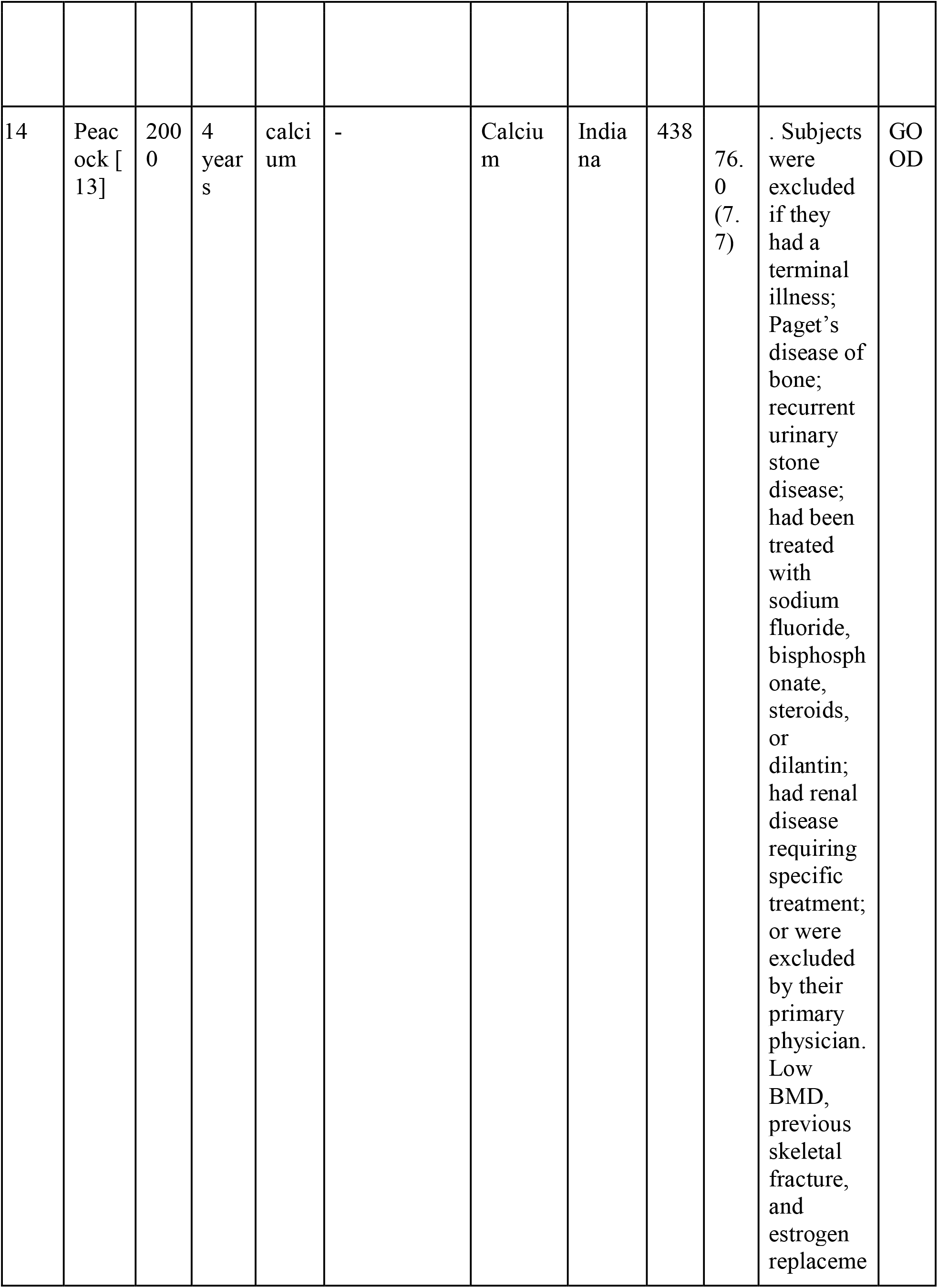
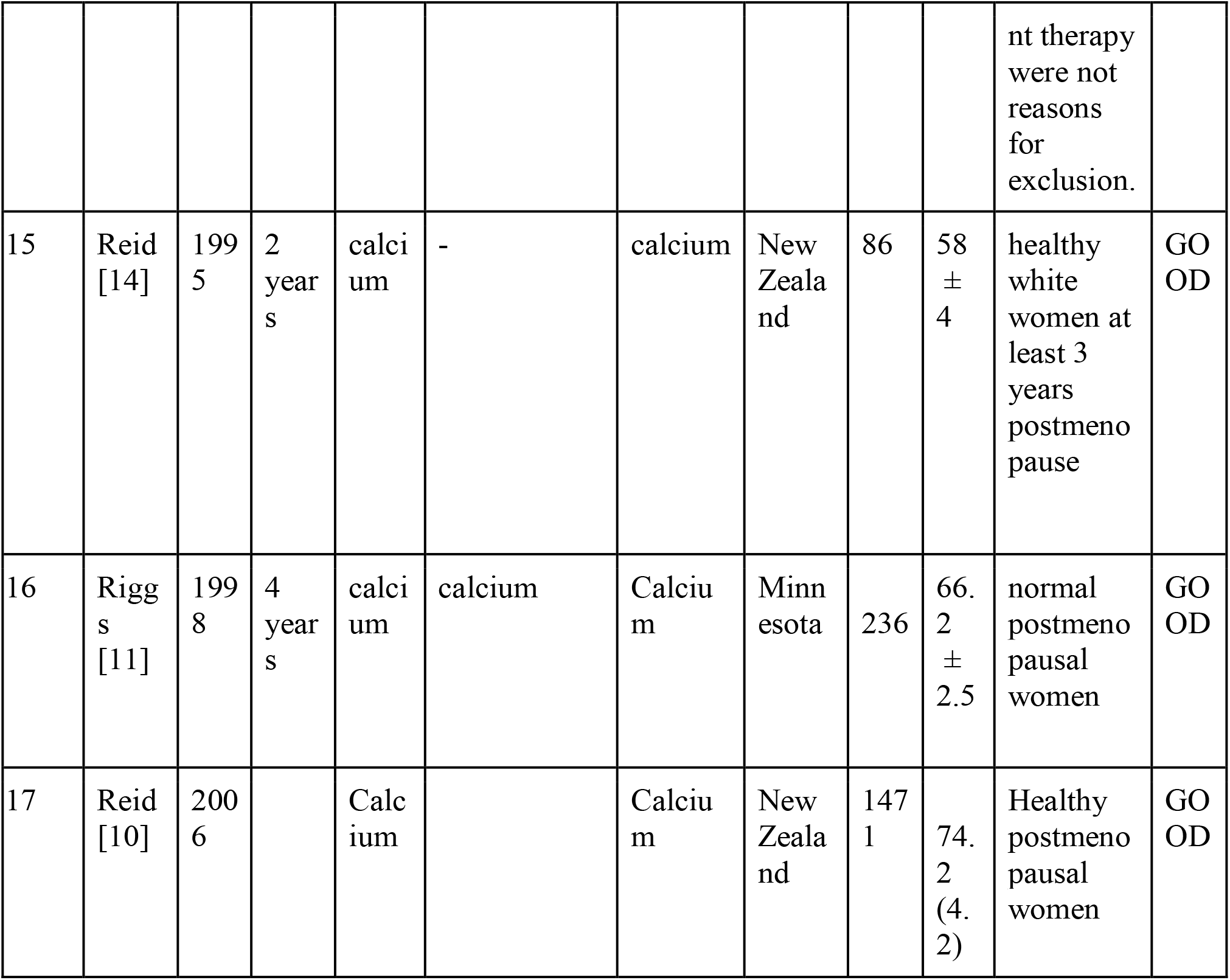

## PRISMA flow chart

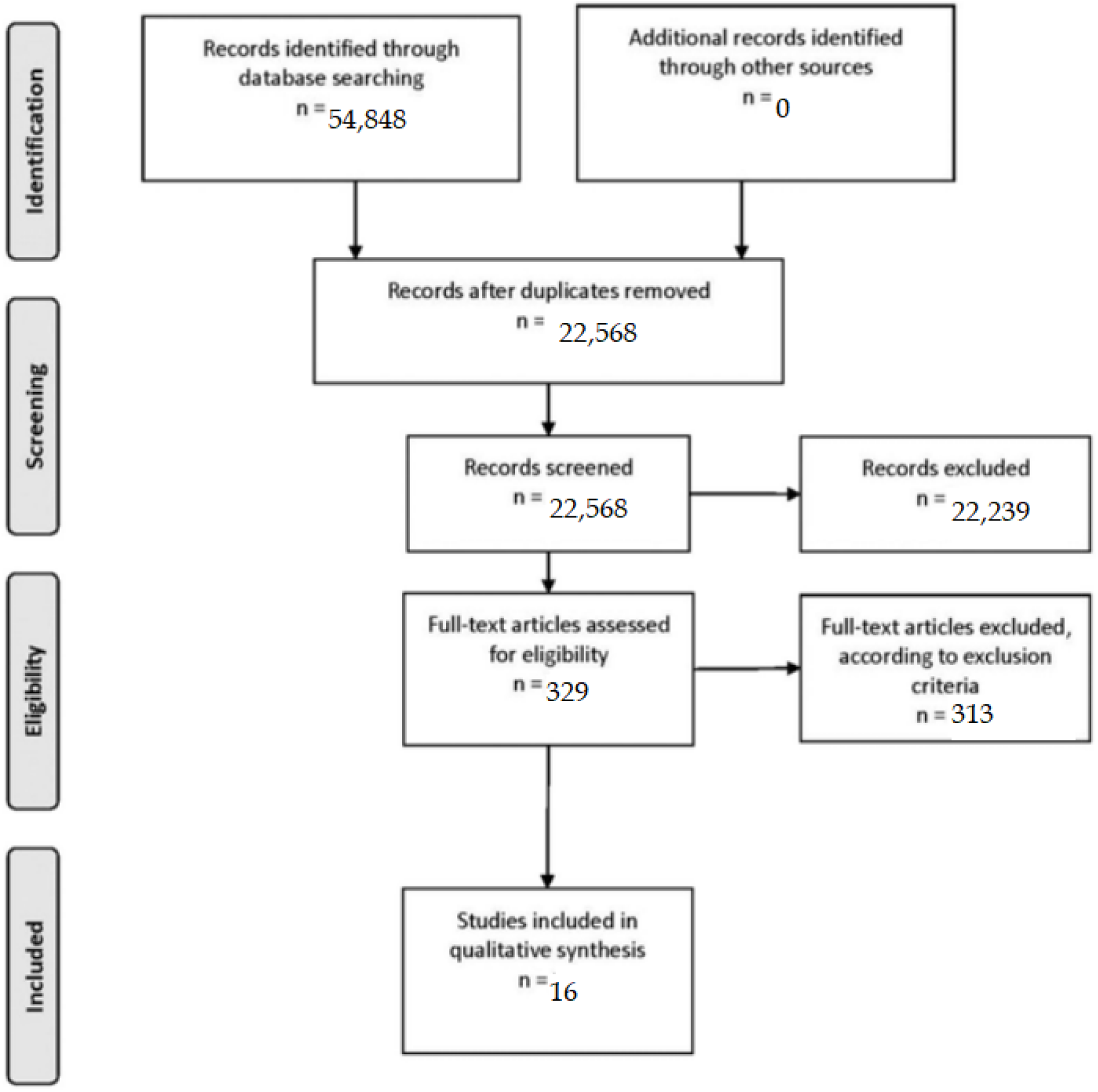

